# Neonatal and Early Childhood Epigenetic Variation Linked to Social and Behavioral Outcomes in Very Preterm Children

**DOI:** 10.64898/2026.02.19.26346629

**Authors:** Priyadarshni Patel, Yijie Huang, Marie Camerota, Casey A. Cragin, Brian S. Carter, Jennifer Check, Jennifer B. Helderman, Julie A. Hofheimer, Elisabeth C. McGowan, Charles R. Neal, T. Michael O’Shea, Steven L. Pastyrnak, Lynne M. Smith, Carmen Marsit, Barry Lester, Todd M. Everson

## Abstract

Very preterm infants (<30 weeks gestation) are at elevated risk for neurodevelopmental and social-behavioral challenges. DNA methylation (DNAm) may provide a biological link between preterm birth and later behavioral outcomes. We examined associations between DNAm profiles at neonatal intensive care unit (NICU) discharge and at age 5 with Social Responsiveness Scale (SRS) scores which measure social communication, social interaction, and repetitive behaviors at age 5, including sex-specific effects, in the Neonatal Neurobehavior and Outcomes in Very Preterm Infants (NOVI) Study. Epigenome-wide buccal DNAm was profiled at NICU discharge (n=218) and at 5 years (n=188). We identified 38 neonatal and 6 age-5 CpG sites associated with SRS scores (all q<0.05) using epigenome-wide association studies (EWAS) at each time point. Several CpGs mapped to genes involved in neurodevelopment including *TCF4*, *KLC4*, *CAP2*, *PTDSS1*, *ADAM12*, *SENP1*, *CHN2*, *SH3D19*, and *ITGA1*, with sex-specific effects observed for CpGs in *CAMTA1* and *GABBR1*. Enriched pathways included neurodevelopment, cytoskeletal regulation, stress-response, and metabolic processes. DNAm patterns during early life, particularly the neonatal period, were associated with social-behavioral development in very preterm children. Findings in key genes such as *TCF4* and *CAMTA1* highlight potential epigenetic mechanisms linking early-life biology to later behavioral challenges.

## INTRODUCTION

Infants born less than 37 weeks’ gestation are classified as preterm^1^ and face an increased risk of long-term developmental and neurobehavioral challenges^2,3^. Those born very preterm (VPT) (< 30 weeks)^1^ are particularly vulnerable, exhibiting higher rates of cognitive, motor, and social difficulties compared to full-term infants^4–6^ Early brain development in these infants is highly sensitive to environmental and physiological stressors, which can shape neurodevelopmental trajectories and influence behavioral outcomes^7,8^.

Epigenetic mechanisms, particularly DNA methylation, may provide a biological framework linking early-life exposures and experiences to neurodevelopment and behavior. Epigenetic mechanisms refer to processes that alter gene expression without altering the underlying DNA sequence ^9^. The neonatal period represents a window of heightened vulnerability for VPT infants. Although lengthy NICU stays are often required to overcome life-threatening conditions, exposure to NICU-related stressors, medical procedures, and environmental fluctuations has been associated with differences in DNA methylation, and these methylation patterns have also been correlated with later social and behavioral outcomes ^10,11^. Epigenome-wide association studies (EWAS) have identified DNA methylation patterns at birth associated with social and behavioral characteristics in very preterm infants ^12^. Early-life DNA methylation may therefore serve as a biomarker of these exposures and their potential impact on developmental trajectories.

By 5 years of age, children have established complex social, cognitive, and behavioral skills^13^. Assessing DNA methylation at this stage allows for the investigation of how early epigenetic patterns persist, evolve, or influence emerging behaviors. Linking neonatal and 5-year methylation profiles with behavioral outcomes provides a longitudinal perspective on the biological mechanisms connecting very preterm birth to social and behavioral development. Additionally, sex differences in autism spectrum disorder are well-documented, with males more frequently diagnosed than females and showing higher levels of autism-related behaviors across social communication and restricted or repetitive domains. In contrast, females often exhibit greater social motivation and compensatory behaviors that may mask underlying difficulties, contributing to under-recognition of ASD-related traits in this group. These differences highlight the importance of examining sex as a biological variable, as associations between early-life epigenetic variation and social responsiveness may differ between males and females ^14^.

The Social Responsiveness Scale (SRS) is a widely used tool for evaluating social abilities and behaviors related to autism spectrum disorder (ASD) in children aged 4–18^15,16^. The updated SRS-2 includes five subscales capturing social communication and interaction (social awareness, cognition, communication, motivation), and restricted/repetitive behaviors (RRB). In children born VPT, SRS scores have been used to study subtle social and behavioral difficulties even in the absence of ASD diagnosis. Social communication difficulties are highly prevalent when measured dimensionally in very preterm populations, suggesting elevated rates of both clinical and subclinical autism related behaviors^17,18^. Early identification of behavioral challenges using tools like the SRS is critical for targeted interventions and optimizing developmental outcomes among VPT children.

Despite the extensive application of EWAS in ASD research^19^, few studies have examined either epigenomic associations with social and behavioral traits in children born very preterm, or associations between DNAm and ASD symptoms at multiple time points. One study has shown that DNA methylation was associated with SRS scores at 5 years of age in children with autism^20^, but overall the literature is sparse. There is a critical need to investigate the neonatal and early childhood biological predictors of autism-related behaviors in the VPT population where these behaviors are more prevalent. To address this need, our study aimed to identify epigenetic features at NICU discharge and at age 5 that were associated with SRS scores measured at age 5, with the goal of advancing the understanding of the biological mechanisms that may underlie the increased risk for social and behavioral problems among children born very preterm. We hypothesize that (1) differential DNA methylation at both time points will associate with SRS Scores, (2) some associations will be sex-specific, and (3) specific CpG sites will correlate with distinct SRS subdomains (e.g., social communication vs. restrictive/repetitive behaviors). Integrating neonatal and 5-year epigenetic data allows for exploration of time-specific and potentially dynamic associations between DNAm and behavioral outcomes.

## METHODS

### Study Population

Participants were drawn from the Neonatal Neurobehavior and Outcomes in Very Preterm Infants (NOVI) Study, a multi-site study of infants born < 30 weeks gestational age (GA). Participants were recruited from nine university affiliated NICUs across six research sites from April 2014 to June 2016. Inclusion criteria were: (1) birth < 30 weeks GA, (2) parental ability to speak English or Spanish, (3) residence within 3 hours of the NICU and follow-up clinic. Exclusion criteria included major congenital anomalies, maternal age < 18 years, cognitive impairment, and death. Parents of eligible infants were approached when infants were 31-32 weeks GA or when survival to discharge was deemed likely by the attending neonatologist. Each site obtained informed consent in line with its institution’s review board. A total of 704 infants were enrolled, and parents of 672 (93%) infants consented to the epigenetics component of NOVI^21^ . Medical and demographic information of participants were collected through medical record abstraction and maternal report at the time of enrollment. Buccal swabs were collected during the week of NICU discharge, and at follow-up visits from 2-5 years of age. The SRS was conducted at the follow-up visit at 5 years of age. Participants were included in this study if they had neonatal DNA methylation data and SRS-2 at 5 years for neonatal EWAS (n=218) and 5-year DNA methylation data and SRS-2 at 5 years for the 5-year EWAS (n=188).

### Behavioral Assessments

The Social Responsiveness Scale, 2^nd^ Edition (SRS-2) was administered to assess autism-related social communication and behavior. The SRS-2 is a caregiver-reported questionnaire that measures severity of social communication deficits and restricted, repetitive behaviors.^22^ It yields a total score as well as subscale scores for specific behavioral domains based on standardized algorithms. The primary outcome variable of this analysis was the total SRS *T*-score, collected during the 5-year visit. Additionally, we examined subscale *T*-scores for the Social Communication and Interaction (SCI) domain and the Restricted and Repetitive Behaviors (RRB) domain to investigate more specific DNAm-behavior associations.

### Genome-wide DNA Methylation Profiling

DNA methylation was evaluated from buccal cell samples collected prior to NICU discharge and again at 5 years of age using the Illumina Infinium MethylationEPIC v1.0 BeadChip. Samples were randomized across plates to minimize batch effects, and raw IDAT files were processed in R using the minfi package. Internal control probes were removed, and detection p-values were used to exclude low-quality samples (>5% probes with p > 0.01) and poorly detected probes (>10% of samples). Functional normalization (preprocessFunnorm) was applied to adjust for technical variation while preserving biological signal, including background, dye-bias, and control probe–based corrections ^23^. Quality was further assessed using EWAStools, and samples with QC flag failures were excluded ^24^. Finally, beta-mixture quantile normalization (BMIQ) was used to correct for probe-type differences ^25^. We used combat to address potential batch effects ^26^. In addition, we excluded CpGs with low variance and outliers defined by 3 times the IQR, and we logit-transformed beta values to M-values prior to analyses. We additionally used CoMeBack to identify co-methylated regions (CMRs), which were then clustered, and principal components analyses were used to represent the expression of each CMR. The neonatal and 5-year DNAm data followed the same processing pipeline but were processed separately as two distinct batches of data. After matching phenotypic data with those who have SRS score at 5 years and DNA methylation at neonatal time point, 218 samples with 496,879 CpGs or CMRs were available for all downstream analysis. For the 5-year time point, 188 samples with 467,230 CpGs or CMRs were available for all downstream analysis.

### Confounder and other Covariates

Based on prior research in this population, neonatal age variables such as GA and age at sample collection were considered as covariates ^27^ . Sex and maternal tobacco use during pregnancy were considered confounders due to the elevated risk for ASD in males and offspring of mothers who smoked during pregnancy ^28,29^. In primary models for both time points, we adjusted for sex. To examine sex-specific associations, sex-by-CpG-specific DNAm was considered as an interaction term in a secondary EWAS. Maternal age greater than 35 years old was also considered a confounding variable due to associations with social and behavioral impairment ^30,31^. Technical batches were not considered as covariates since these were already accounted for using ComBat^32^. We also performed surrogate variable analysis to address residual technical or unmeasured confounders.

Cellular heterogeneity was estimated from a reference panel ^33^. All models included the proportion of epithelial cells as a covariate. Primary caregiver education (< high school diploma or equivalent diploma) was also considered as a covariate based on previous research ^27^. Last, since enrollment into NOVI occurred at six different sites, we also controlled for site in all analyses.

### Statistical Analyses

All statistical analyses were performed in R version 4.4.2. For both time points, we first performed surrogate variable analysis (SVA) to identify and control for residual and unknown confounding factors via the sva package ^34^. Demographic information for the samples, including sex, tobacco use during pregnancy, and participation site, was summarized using counts and proportions of the total participants. Continuous variables, such as age at buccal sample collection and gestational age at birth, were reported as means with standard deviations.

Tabulations of basic demographic information was generated by the gtsummary ^35^ package. For the EWAS, SRS total scores were regressed on CpG m-values via robust linear regression from the MASS ^36^ package, with robust standard errors estimated through the sandwich ^37^ package (clustering by family ID), to identify differentially methylated CpGs at neonatal and 5 year time point. All models were adjusted for potential confounding factors and surrogate variables described above. A secondary EWAS was performed to explore sex-specific effects, by including CpG-by-sex cross-product interaction terms in the model. We used FDR correction of p-values to mitigate the potential for false positives and considered those with q-values < 0.05 to be statistically significant. Visualization of sex-specific associations were generated from visreg ^38^ package, which allowed visualization of sex-specific differentially methylated CpGs while controlling for covariates. We used QQ plots and genomic inflation factor (λ), obtained from qqman package ^39^ to assess potential inflation in all EWAS models. Volcano and Manhattan plots were created by EnhancedVolcano ^40^ and CMplot package ^41^. Gene annotations were obtained from the Illumina’s EPIC methylation arrays in Human Genome version 19. We performed pathway enrichment analysis based on GO and KEGG databases with the methylGSA package ^42–45^. We further did secondary analysis on SRS subscale scores using the most significant CpGs identified in the EWAS models to identify subscale-specific effect sizes for each CpGs which were then visualized with upset plots.

We then explored relevant traits previously associated with the top hits. For each CpG identified in any of the EWAS models, we retrieved the traits linked to genes annotated to these CpGs in prior GWAS studies via the GWAS catalog (https://www.ebi.ac.uk/gwas/docs/file-downloads). To focus on the most robust associations, we highlighted traits supported by at least 10 studies.

## Results Demographics

Demographic characteristics of the study sample at the neonatal and 5-year timepoints are summarized in Table 1. At the neonatal timepoint, the sample included 218 children (119 males, 99 females) with a mean gestational age of 27.10 weeks (SD ± 0.13).

**Table 1.** Demographic.

| Table 1 : Demographic |  |  |  |
| --- | --- | --- | --- |
| **Neonatal** |  |  |  |
| **Variable** | **Overall**<br>**(218)** | **Male**<br>**(119)** | **Female**<br>**(99)** |
| Gestational Age at birth (weeks) | 27.10 ±<br>0.13 | 27.20 ±<br>0.17 | 26.97 ±<br>0.19 |
| Gestational age at sample collection<br>(weeks) | 39.83 ±<br>0.27 | 39.39 ±<br>0.32 | 40.35 ±<br>0.45 |
| Hollingshead SES |  |  |  |
| <HS/GED | 21 (10%) | 15 (13%) | 6 (6%) |
| Maternal age >35 at childbirth |  |  |  |
| Yes | 54 (25%) | 28 (24%) | 26 (26%) |
| Maternal Tobbaco Use |  |  |  |
| Yes | 24 (11%) | 11 (9%) | 13 (13%) |
| **60 months** |  |  |  |
|  | **Overall**<br>**(188)** | **Male (99)** | **Female**<br>**(89)** |
| Gestational Age at birth (weeks) | 26.93 ±<br>1.93 | 27.04 ±<br>1.87 | 26.80 ±<br>1.99 |
| Age at sample collection (months) | 68.65 ±<br>2.98 | 68.89 ±<br>2.93 | 68.40 ±<br>3.04 |
| Hollingshead SES |  |  |  |
| <HS/GED | 17 (9%) | 11 (11%) | 6 (6.7%) |
| Maternal age >35 at childbirth |  |  |  |
| Yes | 55 (29%) | 28 (28%) | 27 (30%) |
| Maternal Tobbaco Use |  |  |  |
| Yes | 17 (9%) | 9 (9%) | 8(9%) |
| Note: Values are shown as mean ± SD for continuous variables and n (%) for categorical variables. P-values reflect comparisons between males and females using t-tests (continuous) or chi-square tests (categorical). HS/GED indicates caregiver education level. SES is based on the Hollingshead Index. “Epithelial Cells (%)” represents the proportion of epithelial cells in the collected samples. |  |  |  |

The average age at neonatal sample collection was approximately 40 weeks GA. Most caregivers (90%) had at least a high school diploma or equivalent level of education. At the 5- year timepoint, data were available for 188 children (99 males, 89 females). The mean GA at birth for the 5-year sample was similar to the neonatal sample (26.93 weeks ± 1.93), and children were on average 68.65 months old at the time of 5-year sample collection. Caregiver educational attainment (91% with HS/GED or higher), maternal age at birth >35 years (29%) and prenatal tobacco exposure (9%) were similar compared to the neonatal sample (Table 1).

### Social Responsiveness Scale (SRS)

The mean SRS total *T*-score measured at age 5-year was 53 (*SD* = 10) (T-scores > 60 indicate clinically elevated symptoms), with a median of 51 (IQR = 11), indicating that, on average, scores were within the normative range. Among the subdomains, the Social Communication and Interaction T-score had a mean of 53.64 (SD = 9.93) with a median of 52 (IQR = 11) while the Restricted and Repetitive Behaviors (RRB) had a mean score of 51.71 (SD= 10.68) with a median of 48 (IQR = 11).

### Neonatal EWAS Findings

When regressing SRS-2 *T-* scores on neonatal DNAm, this epigenome-wide association study (EWAS) yielded an inflation factor λ = 0.99 indicating no inflation. We found that DNA methylation at 38 CpG sites was associated with the SRS scores (q <5%; Table 2A). Of these, there were 12 positive associations (i.e., higher DNAm associated with higher SRS-2 scores) and 26 negative associations (i.e., lower DNAm associated with higher SRS-2 scores). Overall, the associations were moderate in magnitude, where differences in DNAm from the 25th to 75th percentile were associated with approximately 6.3-to-6.8-point differences in the SRS-2 T-score. Figure 1 shows the genomic organization of our findings with a Manhattan plot, and the distribution of effects sizes and p-values via volcano plots. We used functional enrichment tests to identify broader pathways or processes that may be represented among the differentially methylated CpGs. After FDR correction (q < .05), we identified four GO processes that were significantly enriched (*phosphatidylinositol phosphate 4-phosphatase activity, G-protein beta-subunit binding, peptide antigen binding, positive regulation of translational initiation*). No significant pathways were identified in KEGG.

**Figure 1.**
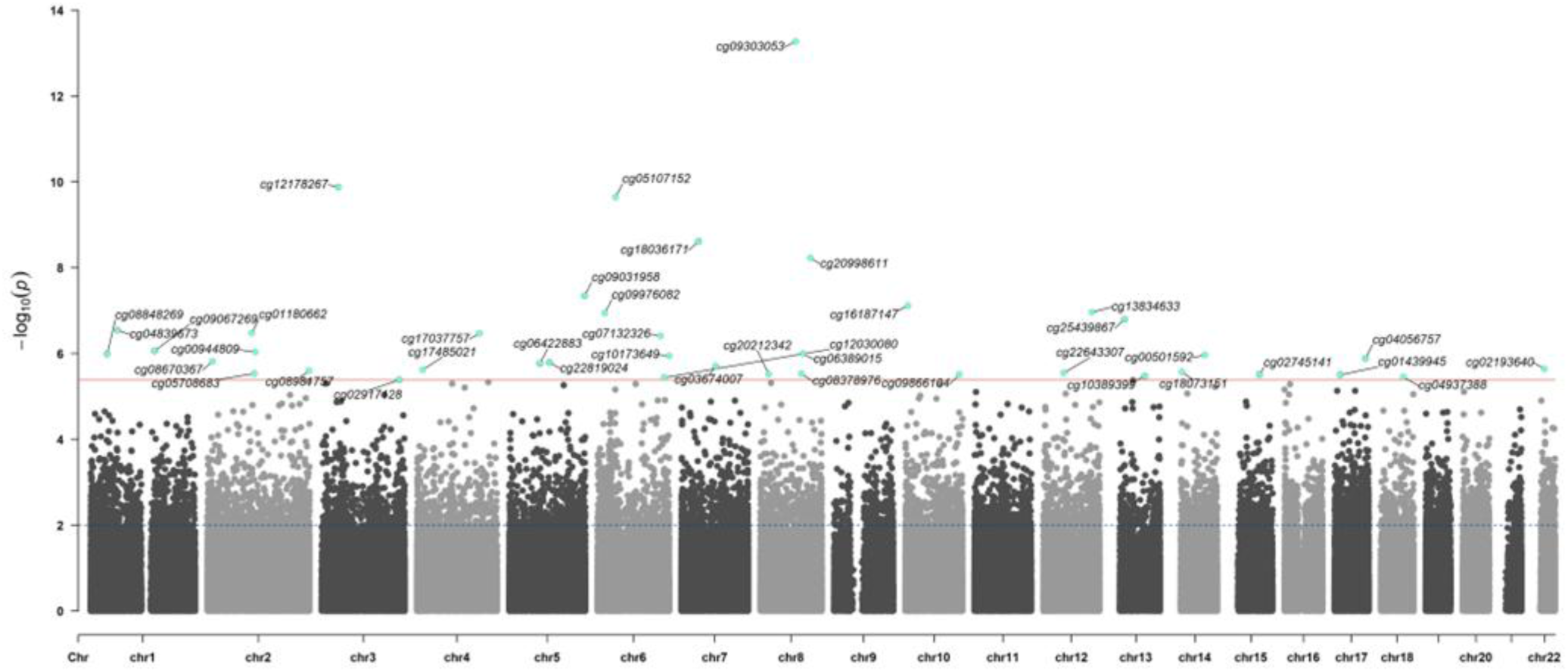
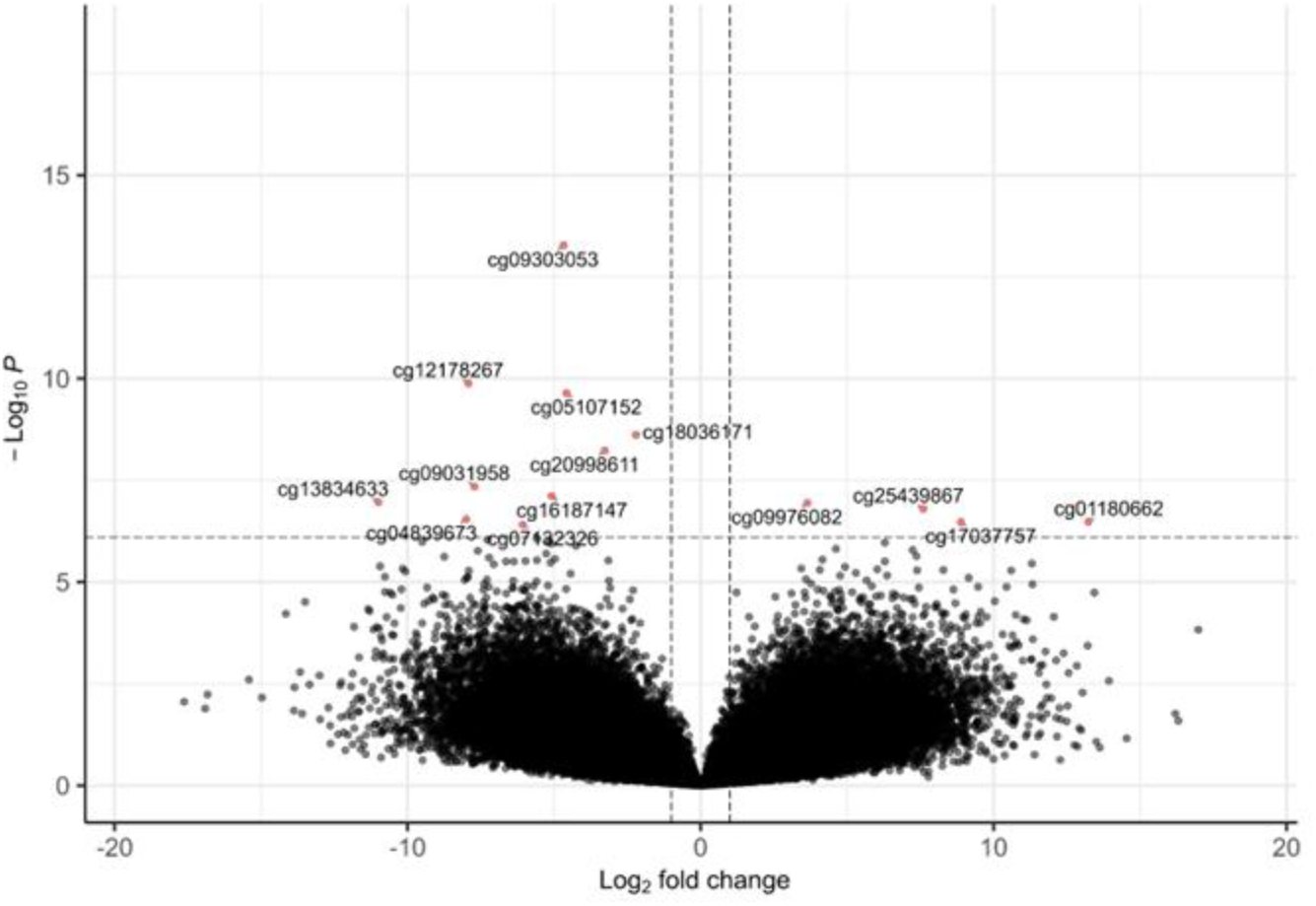
Manhattan and Volcano plot of neonatal epigenetic loci associated with 5-year SRS scores.

**Table 2.**
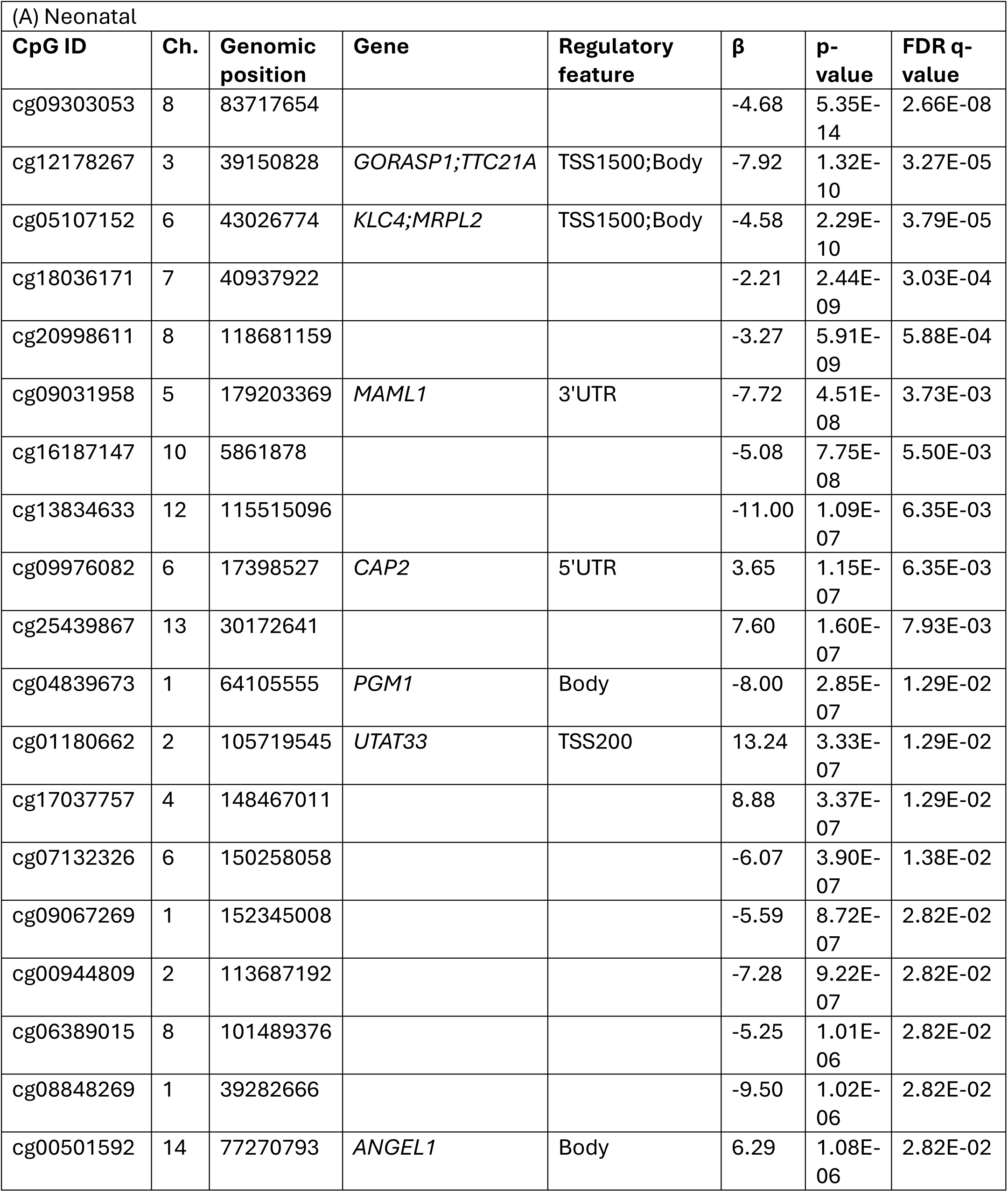
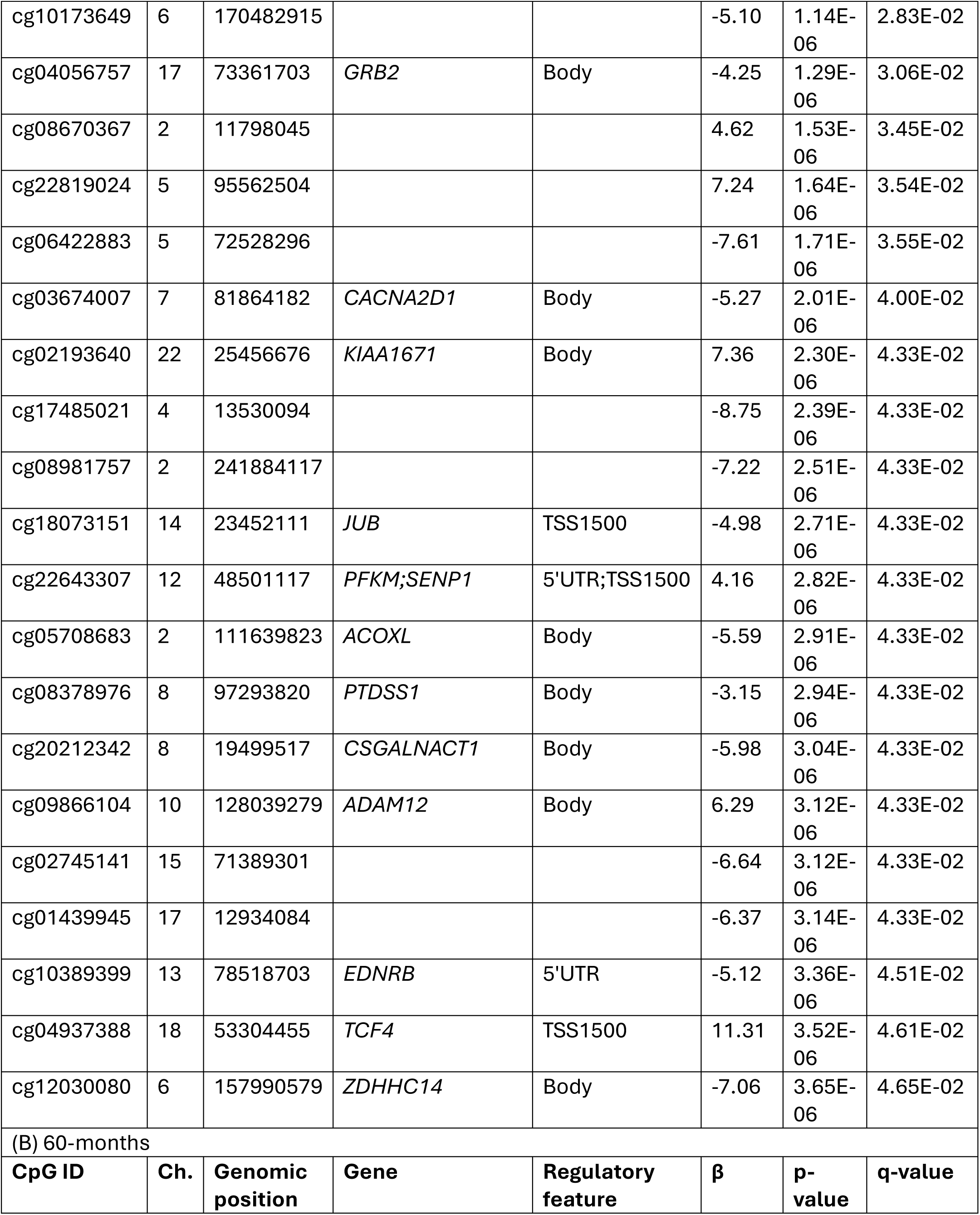
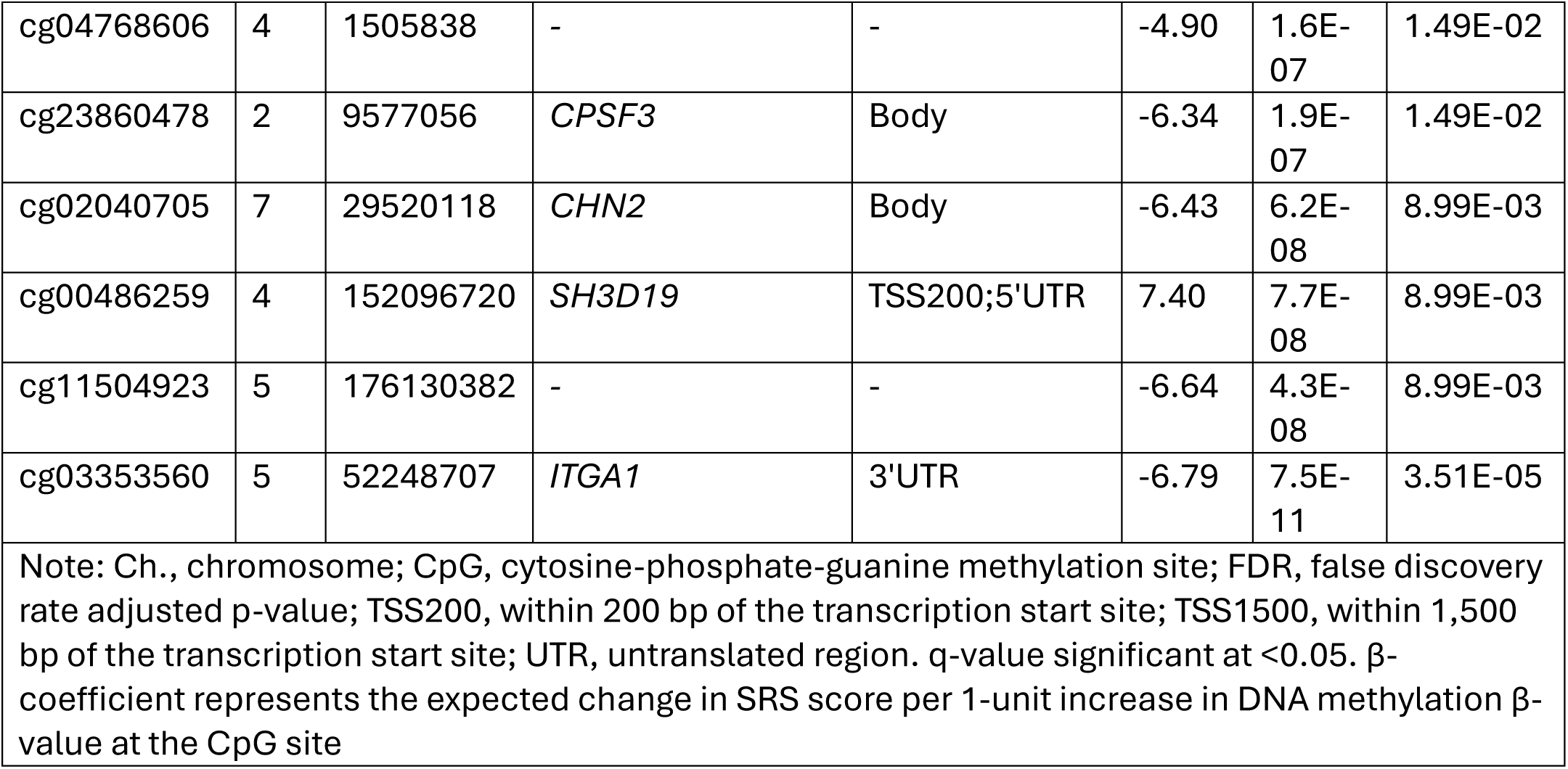
Epigenome-wide association study results for statistically significant CpG sites (FDR<5%) at (A) Neonatal period (B) 60 months.

For each of the identified CpGs, we examined traits that have previously been associated with the genes annotated to these CpGs (Supplemental Table 1A). There were several relevant phenotypes and traits associated with the genes annotated to the significant CpG sites from prior GWAS across three major domains: behavioral/neuropsychiatric (autism spectrum disorder, schizophrenia, depression, neuroticism, and educational attainment), sleep-related (insomnia and short sleep duration), and metabolic health (body mass index, metabolic syndrome, and triglyceride levels). Overall, the genes annotated to CpG sites associated with SRS scores were enriched for biological pathways related to neurodevelopment, sleep regulation, and metabolic processes.

As a secondary analysis, we included CpG-by-sex interaction terms in the EWAS to determine whether there were sex-specific epigenetic predictors of SRS scores in children. DNA methylation at 6 CpG sites exhibited significant sex-interaction terms (FDR q-values < 0.05; Table 3A). Supplemental Figure 1 demonstrates the genomic organization of the sex-specific associations and distribution of effects via Manhattan and volcano plots. For two of the sex-specific CpGs (cg26893022 and cg22827496), the associations appear to be driven by outliers’ values, while 5 CpGs exhibit more robust and apparent sexually dimorphic associations between DNAm and SRS-2 scores (Figure 3; Panels 1-7).

**Figure 2.**
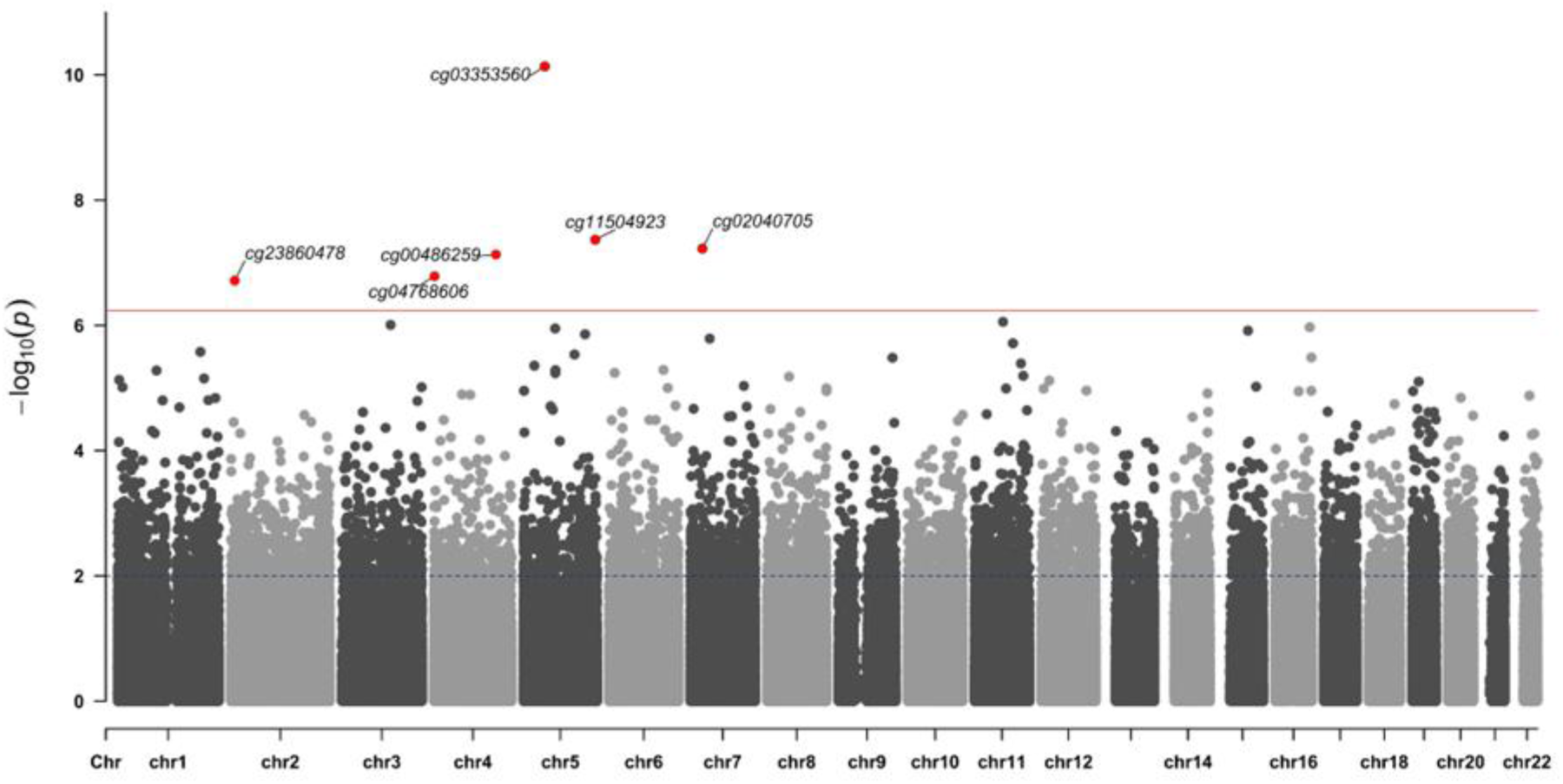
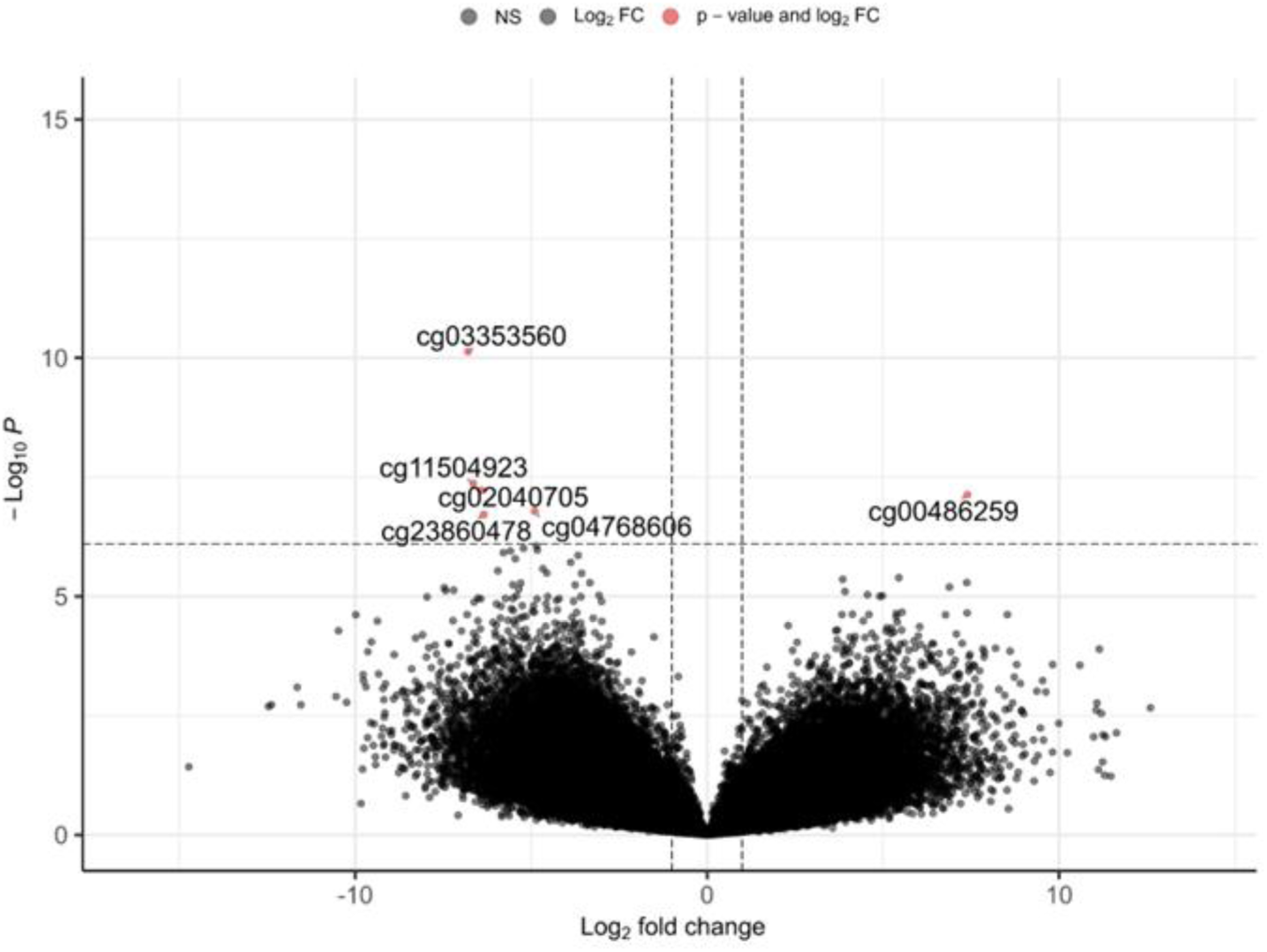
Manhattan plot and Volcano plot of 5-year epigenetic loci associated with 5-year SRS scores.

**Figure 3.**
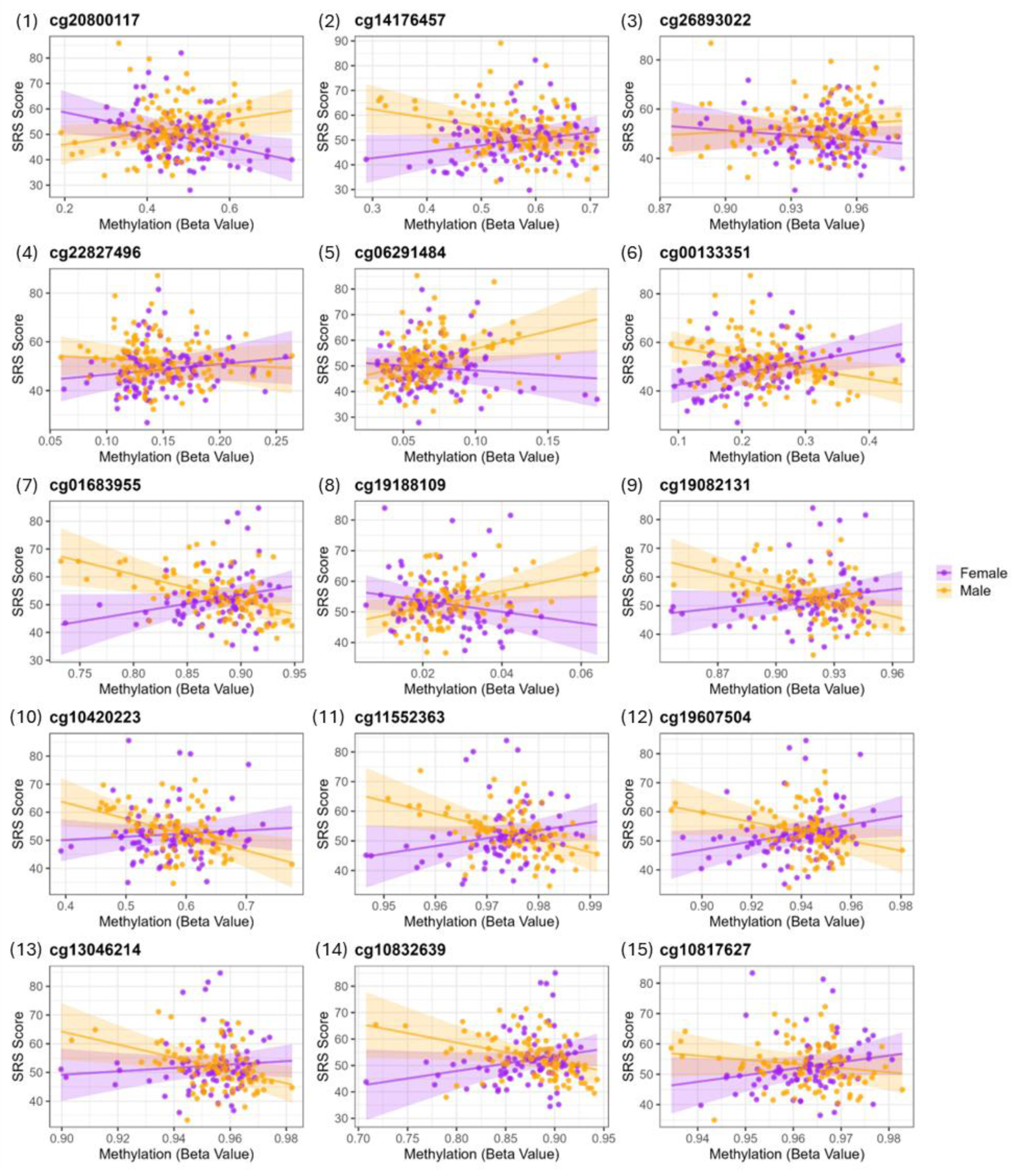
Sex-specific CpG associations with SRS scores in the neonatal (Panels 1–6) and 5-year (Panels 7–15) EWAS. Panels shows CpG sites with significant sex interactions, highlighting differential methylation effects on SRS outcomes in males (yellow) and females (purple)

**Table 3.** Sex interaction epigenome-wide association study results for statistically significant CpG sites (FDR<5%) at (A) Neonatal period (B) 60 months.

| CpG ID | Ch | Genomic position | Gene | Regulatory feature | $\beta$ | p-value | FDR q-value |
| --- | --- | --- | --- | --- | --- | --- | --- |
| cg20800117 | 1 | 7308366 | *CAMTA1* | Body | 11.25 | 4.42E-09 | 2.20E-03 |
| cg14176457 | 3 | 69698369 |  |  | -13.62 | 1.25E-07 | 2.07E-02 |
| cg26893022 | 4 | 38511077 | *LINC01258* | Body | 8.02 | 3.52E-07 | 2.89E-02 |
| cg22827496 | 6 | 12014450 | *HIVEP1* | 5'UTR | -7.35 | 2.95E-07 | 2.89E-02 |
| cg06291484 | 11 | 94245547 |  |  | 11.70 | 7.97E-08 | 1.98E-02 |
| cg00133351 | 20 | 37070585 |  |  | -11.99 | 4.08E-07 | 2.89E-02 |

(B) 60-months
| CpG ID | Ch | Genomic position | Gene | Regulatory feature | $\beta$ | p-value | FDR q-value |
| --- | --- | --- | --- | --- | --- | --- | --- |
| cg01683955 | 1 | 100251901 |  |  | -12.1053 | 9.05E-07 | 0.046957 |
| cg19188109 | 3 | 138327414 | *FAIM* | TSS1500;TSS2000 | 10.20768 | 2.94E-07 | 0.034342 |
| cg19082131 | 4 | 2363153 | *ZFYVE28* | Body | -12.5427 | 8.43E-07 | 0.046957 |
| cg10420223 | 6 | 29526561 | *UBD, GABBR1, OR2I1P* | Body | -13.0522 | 5.22E-07 | 0.046957 |
| cg11552363 | 8 | 141059899 | *TRAPPC9* | Body | -12.1273 | 7.56E-07 | 0.046957 |
| cg13046214 | 12 | 133401908 | *GOLGA, CHFR* | 5'UTR | -11.4413 | 1.27E-07 | 0.02556 |
| cg19607504 | 12 | 117794293 | *NOS1* | 5'UTR | -13.7341 | 2.26E-08 | 0.010551 |
| cg10832639 | 14 | 94850965 | *SERPINA*<br>1 | 5'UTR | -<br>11.0542 | 8.65<br>E-07 | 0.04695<br>7 |
| cg10817627 | 20 | 7075613 |  | - | -<br>12.9333 | 1.64<br>E-07 | 0.02556 |

Note: Ch., chromosome; CpG, cytosine-phosphate-guanine methylation site; FDR, false discovery rate adjusted p-value; TSS200, within 200 bp of the transcription start site; TSS1500, within 1,500 bp of the transcription start site; UTR, untranslated region. q-value significant at <0.05. $\beta$ -coefficient represents the expected change in SRS score per 1-unit increase in DNA methylation $\beta$ -value at the CpG site.

Functional enrichment tests identified one KEGG pathway (long-term depression) that was significant among the sex-specific CpGs. No GO process was identified. The GWAS traits (Supplement Table 2A) that appeared most frequently among top signals for these sex-specific CpGs were related to growth (body mass index, height) and behavioral domains (neuroticism, educational attainment, and externalizing behavior).

### 5-year EWAS Findings

When regressing SRS-2 total scores cross-sectionally on the 5-year DNAm data, this EWAS yielded an inflation factor λ = 0.91 again indicating no inflation. DNA methylation at 6 CpG sites was associated with SRS-2 T-scores (q < .05; Table 2B). Of these, there was 1 positive association and 5 negative associations. Overall, the associations were moderate in magnitude where comparing the 25th to 75th percentile of DNAm was associated with approximately a 6.4-to-6.7-point changes in the SRS-2 T-score. Manhattan and volcano plots of 5-year EWAS results are presented in Figure 2. We identified significantly enriched (FDR q < 0.05) GO processes (*P-type calcium transporter activity, fatty acid elongase activity, integrin comples*), while no significant pathways were identified in KEGG. The GWAS traits for the genes annotated to these CpGs included autism spectrum disorder, general cognitive ability and educational attainment (Supplemental Table 1B).

Our secondary cross-sectional EWAS tested for sex-specific effects, identifying DNA methylation at 9 CpG sites with significant CpG-by-sex interactions (q-values < 0.05; Table 3B) that were distributed across the genome (Supplemental Figure 2). For all 9 of these CpGs, the sexual dimorphism of the associations was evident with DNAm exhibiting opposing directions of association with SRS scores for males and females (Figure 3; Panels 8 - 16). We identified four significant (FDR q-values < 0.05) KEGG pathways (*antigen processing and presentation, arginine and proline metabolism, Type 1 diabetes mellitus, autoimmune thyroid disease*), but no significant GO processes. For the identified CpGs, the most common GWAS traits (Supplementary Table 2B) included metabolic and growth-related factors (body mass index, height, type 2 diabetes), cardiovascular regulation (electrocardiogram morphology, QT interval, blood pressure), and behavioral/neuropsychiatric traits (autism spectrum disorder or schizophrenia, social communication problems, insomnia, and cognitive function).

### SRS2 Subscale Analysis

We examined the top hits from our neonatal and 5-year EWAS with the SCI and RRB domain *T-scores* to explore domain-level associations (Supplemental Table 3). At the neonatal time point, 18 of 38 CpGs showed significant associations with the SCI domain, and 11 CpGs with the RRB domain. At the 5-year time point, 3 CpGs and 6 CpGs were significantly associated with the RRB and SCI domains, respectively. The CpG sites associated with social communication and repetitive behavioral traits showed overlapping patterns of methylation, indicating shared epigenetic signatures across social-behavioral domains.

## Discussion

The purpose of this study was to identify DNAm predictors of autism-related behaviors in children born very preterm. We found 38 CpGs at the neonatal time point and 6 CpGs at the 5-year time point that were significantly associated with 5-year SRS scores, indicating stronger overall signal in the neonatal epigenetic data. Prior EWAS examining autism-related behaviors, including the SRS-2, remain limited and differ both in tissue type and study design. A case-control study in whole blood in children identified more localized findings, including a differentially methylated region near *ZFP57* associated with SRS scores ^20^. Another meta-analysis found that DNAm signals vary by tissue, developmental stage, and analytic approach, with different tissues showing largely distinct, but some overlapping, autism related pathways ^19^. Additionally, Mouat et al. ^46^ demonstrated sex-specific DNA methylation signatures in newborn blood, showing that males and females diagnosed with ASD exhibit different DNA methylation patterns and enrichment results. Our research focused on a very preterm high-risk population with readily accessible buccal tissue swabs to identify DNAm–behavior associations at multiple developmental stages, as well as some sex-specific relationships.

Our findings for the neonatal EWAS in particular, identified several epigenetic loci annotated to genes with known roles in neurodevelopment, behavior, and neurodegeneration, including *TCF4*, *KLC4*, *CAP2*, *PTDSS1*, *ADAM12*, and *SENP1*. Of particular note is the gene *transcription factor 4* (*TCF4*; annotated to cg04937388), where neonates with higher DNAm were more likely to have higher SRS-2 total scores at 5 years of age. Pitt-Hopkins syndrome (PTHS), a rare genetic condition that is considered an autism spectrum disorder and linked to severe language impairment and developmental delay, is caused by human haploinsufficiency of the transcription factor *TCF4* ^49^. Others ^50^ have shown that *TCF4* is expressed during peak neurogenesis, and its loss causes brain malformations along with widespread disruption of genes involved in neuronal development and neurodevelopmental disorders. Interestingly, our identified CpG lies in the transcription start site upstream of *TCF4*, and higher methylation in promoter regions is typically associated with repression of expression. While we did not have the appropriate data to specifically test whether high DNAm at cg04937388 represses *TCF4* expression, our observation of increased methylation theoretically aligns with the above studies where loss, repression, or dysfunction of *TCF4* was linked to adverse neurodevelopmental outcomes including a rare genetic forms of ASD. Our identification of higher DNAm upstream of *TCF4* highlights the biological plausibility of our findings, and indicates that perturbed DNAm, possibly epigenetic repression in the promoter of *TCF4* in early development may be related to autism-like behaviors later in childhood.

In line with this, we also observed a significant association at a CpG site (cg05107152) annotated to *KLC4*, a kinesin light chain subunit involved in neuronal cargo transport where higher methylation was associated with lower SRS scores. *KLC4* is important for establishing neuronal morphology through its role in axon branching, microtubule regulation, and endosomal trafficking ^51^. Notably, experimental work in zebrafish models has shown that loss of *KLC4* disrupts axonal compartmentalization and leads to abnormal branching patterns, as well as behavioral alterations including heightened stress sensitivity ^51^. These findings provide mechanistic insight into how variation in *KLC4* regulation could contribute to altered neuronal connectivity and behavioral phenotypes, in particular among children born preterm. Interestingly, the identified CpG (cg05107152) lies within the transcription start site upstream of *KLC4,* and within the first intron of *MRPL2,* a mitochondrial ribosomal protein which is important in mitochondrial function and may be involved in cellular energy synthesis ^52^. Both *KLC4* and *MRPL2* are highly expressed in brain tissue. Methylation changes at this site could plausibly be related to specific neuronal development pathways or have broader impact on cellular energy regulation, providing biological plausibility for its potential link to neurodevelopmental outcomes.

Another study ^53^ examining DNA methylation profiles from buccal swabs in children with autism spectrum, identified significant differentially methylated patterns in *CAP2* gene, also annotated to one of our top hits. This is particularly relevant because *CAP2* encodes cyclase-associated actin cytoskeleton regulatory protein 2, which is highly expressed in the brain ^54,55^, suggesting that altered epigenetic regulation of *CAP2* may similarly impact neuronal structure and function. The *PTDSS1* gene (cg08378976) also emerged among our top neonatal EWAS results. In a Lebanese population-based copy number variation (CNV) study ^56^, *PTDSS1* was identified alongside several others as a novel autism susceptibility gene, marking it as a potential contributor to ASD likelihood through genomic alterations. This convergence of CNV and DNAm evidence at *PTDSS1* underscores its potential role at the intersection of genetic and epigenetic mechanisms affecting neurodevelopment. Another gene from our neonatal EWAS, *ADAM12* belongs to a large family of multidomain metalloprotease with both cell-binding and proteolytic functions ^57^. Within the human and rat CNS, *ADAM12* is primarily expressed in oligodendrocytes of both white and gray matter, and is also detectable in astrocytes, neurons, and endothelial cells ^58^. While its role in the healthy brain remains unclear, increasing evidence links *ADAM12* dysregulation to several neurological and psychiatric conditions, including brain tumors ^59^, Alzheimer’s disease ^60^, schizophrenia ^61^, autism spectrum disorder ^62^, and bipolar disorder ^63^. Finally, recent research has also identified *SENP1* as a significant potential gene for ASD. The whole exome sequencing of individuals with ASD identified a novel heterozygous protein-truncating mutation in *SENP1* ^64^. Mouse models with *SENP1* haploinsufficiency show significant social abnormalities and repetitive habits, while mutations in *Senp1* are linked to serious neurological diseases ^65^.

Taken together, our neonatal EWAS identified multiple CpGs within key neurodevelopmental genes such as *TCF4*, *KLC4*, *CAP2*, *PTDSS1*, *ADAM12*, and *SENP1,* providing evidence that DNAm variation measured as early as NICU discharge may capture perturbed signals in pathways fundamental to neurodevelopment. These findings raise the possibility that epigenetic alterations detectable in the neonatal period contribute to the emergence of autism-related behaviors observed years later. Such observations suggest that neonatal DNAm profiling could offer an early window into long-term neurodevelopmental outcomes in very preterm infants. In turn, they underscore the importance of further work to establish how these early epigenetic signatures might inform risk stratification and guide the timely delivery of interventions aimed at improving developmental outcomes.

While the neonatal DNAm findings highlight early-life epigenetic influences on neurodevelopment, our EWAS at 5-year provides complementary insight into DNAm patterns closer to the time when autism-like behaviors are measured. These cross-sectional associations at 5-year may reflect influences on ongoing epigenetic regulation throughout early childhood and how these are related to social and behavioral outcomes. Similar to genes from the neonatal EWAS, the genes annotated to our 5-year EWAS were involved in neurodevelopment and brain function. One of the notable CpG sites (cg02040705) is annotated to *CHN2*, a gene implicated in neurological development. Elia et al. ^66^ found that rare inherited structural variants in ADHD patients were enriched in genes associated with neurodevelopmental disorders, including *CHN2*. We additionally identified *ITGA1* (cg03353560), which has previously been identified as a susceptibility gene for ADHD ^69^. Overall, our 5-year EWAS identified a smaller number of CpGs than the neonatal EWAS, yet our top hits were similarly biologically significant in that they annotated to genes with known neurodevelopmental functions. These results indicate that these loci may be under differential epigenetic regulation during the period when autism-related behaviors are commonly expressed.

We also evaluated whether sex moderates the association between DNA methylation and SRS-scores. Notably, we did identify multiple CpGs that appear to exhibit sex-specific associations. Again, several of these CpGs were annotated to genes involved in neurodevelopment, highlighting their potential functional relevance to autism-like behaviors.

At the neonatal timepoint, the most significant sex-interaction signal was at cg20800117, annotated to *CAMTA1*. Genetic evidence strongly supports the role of *CAMTA1* in brain function and neurodevelopment. Intragenic rearrangements of *CAMTA1* have been described, with affected individuals presenting with a range of phenotypes including intellectual disability, developmental delay, behavioral difficulties, and cerebellar abnormalities ^70,71^. Another cohort has reinforced these associations, highlighting consistent clinical features such as motor and speech delay, hypotonia, intellectual disability, cerebellar dysfunction, and feeding difficulties ^72^. In our cohort, males displayed significant positive association and females had negative association between cg20800117 with the SRS scores. Interestingly, *CAMTA1* has been implicated to have sex-specific associations with other phenotypes. For instance, SNPs in and near *CAMTA1* have been associated with increased risk of chronic obstructive pulmonary disease, but only among females ^73^ and a recent pre-print found that females tend to have higher expression of *CAMTA1*, and that this sex-biased expression may offer some protection against neurodevelopmental disorders among females ^74^. Taken together, these findings implicate *CAMTA1* as a gene essential for neurodevelopmental processes, with alterations linked to a broad spectrum of cognitive and behavioral impairments, and emerging evidence that it’s activity may be differentially associated with outcomes for males and females.

At the 5-year timepoint, one of the notable sex-specific findings was cg10420223 within the GABA-B receptor subunit 1 gene *(GABBR1).* Previous work ^75^ has shown that *GABBR1* methylation in cord blood is associated with maternal pregnancy anxiety, with stronger effects observed in male newborns. A top differentially methylated region within *GABBR1* was significantly correlated with infant cortisol reactivity at 4 months of age, pointing to a role for this locus in programming stress-response pathways ^75^. We observed significant positive associations between cg10420223 and SRS scores with males and negative associations with females. *GABBR1* has been implicated to have sexually dimorphic roles elsewhere. Others have shown that *GABBR1* is a sex-specific target of the transcription factor *Nkx6.1*, mediating the regulation of astrocyte morphology and cholinergic synapse formation differently between males and females in the spinal cord ^76^.

Our observed sex-specific associations involving *GABBR1* add to the evidence that this gene may play a role in sex-differences in neurodevelopment. Interestingly, *TRAPPC9* also emerged in our analyses, and mutations in this gene have been linked to intellectual disability, developmental delay, and autism spectrum disorder ^77,78^. An experimental mouse study found that *TRAPPC9* deficiency was linked to smaller brain size, behavioral impairments, and increased weight gain, and that female mice also exhibited increased fat mass accumulation and elevated lipid profiles ^79^.

Our sex-interaction EWAS findings across both neonatal and childhood timepoints suggest that some of the epigenetic influences on autism-like behaviors may differ between males and females. These sex differences could reflect biological variation in how DNA methylation patterns are established and maintained, and how different exposures and experience during early childhood may influence epigenetic regulation, and impact brain development and behavior. Males may be more vulnerable to showing autism-like behaviors, while females may show different or more subtle patterns, consistent with the known differences in autism risk between sexes ^14^.

Together, these results highlight the importance of considering sex as a key factor when studying the epigenetic pathways and neurodevelopment. Additionally, our overall findings suggest that both early (neonatal) and later (childhood) DNA methylation patterns are associated with autism-like behaviors. These findings highlight multiple potential windows of developmental sensitivity, as well as areas potentially amenable to early targeted interventions. These epigenetic patterns likely reflect cumulative contributions to neurodevelopment, emphasizing the complex biology underlying social behavior.

Beyond the established neurodevelopmental functions and associations of the highlighted genes above, the loci identified in our EWAS likely have broad implications for health and development. Our search of the GWAS catalog revealed that our SRS-associated genes are involved in diverse biological systems, encompassing metabolic regulation (e.g., body mass index, lipid and fatty acid metabolism, type 2 diabetes, glycated hemoglobin), hematologic and immune pathways (e.g., red blood cell and monocyte counts), behavioral and psychiatric phenotypes (e.g., insomnia, depression, neuroticism, schizophrenia, autism spectrum disorder, ADHD), and cognitive and educational attainment. These patterns align with growing evidence that children with autism-related behavioral differences often experience broader physiological challenges, including alterations in immune function and cardiometabolic health ^80 81^. This is reflected in the methylation signatures that we identified here, which appear to capture systemic biological processes that extend beyond behavior alone.

We note that our study has several limitations which should be considered when interpreting our findings. First, our sample size is moderate, and thus we may have missed very small associations or may be prone to identifying false positive associations. However, the biological plausibility of our identified CpGs and genes, and prior evidence for sexually dimorphic roles, provide supporting evidence that our findings are robust. Second, DNA methylation was measured in buccal cells, which may not fully capture epigenetic changes in brain tissues, though prior studies suggest moderate correlations between buccal and brain methylation for some loci ^82^. Third, while we adjusted for known confounders as well as unmeasured confounding with SVA, residual environmental or genetic factors could still influence the observed associations. Finally, our cohort included children born very preterm, a group known to be at elevated risk for social impairment and ASD. Thus, our findings may not be generalizable to other children such as those born at term or with other risk factors for ASD. These limitations provide important context needed to interpretate our findings. Overall, we report a robust set of epigenetic features and their associated genes, that are predictive of or associated with autism-like behaviors, many of which have known roles in neurodevelopment, and some of which may have influence neurodevelopment in sex-specific ways.

## Conclusion

In conclusion, our study demonstrates that DNA methylation patterns in the neonatal and early childhood periods have specific associations with autism-like behaviors in children born very preterm. We also demonstrate there may be sex-specific biological pathways underlying emergence of social impairment. Our findings provide new insight into how early-life epigenetic variation could contribute to neurodevelopment and social behavior, suggesting that patterns of DNAm could be used for early identification of children at increased likelihood for ASD and related behaviors. Future studies with larger, diverse cohorts and integration of multi-omics data will be important to validate and extend these findings.

## Data Availability

All data produced in the present study will be available after publication

---

**Supplement table 1.** GWAS Trait frequency in (A) neonatal (B) 60 months.

| **Supplement table 1. GWAS Trait frequency in (A) neonatal (B) 60 months** |  |
| --- | --- |
| **(A) Neonatal** |  |
| **Trait** | **n** |
| Height | 68 |
| Monounsaturated fatty acids to total fatty acids percentage | 40 |
| Polyunsaturated fatty acids to monounsaturated fatty acids ratio | 34 |
| Body mass index | 33 |
| Insomnia | 32 |
| Multi-trait sex score | 24 |
| Type 2 diabetes | 24 |
| Monounsaturated fatty acid levels | 22 |
| Schizophrenia | 22 |
| Monocyte count | 19 |
| Educational attainment | 18 |
| Omega-6 fatty acids to total fatty acids percentage | 18 |
| Red blood cell count | 18 |
| Polyunsaturated fatty acids to total fatty acids percentage | 17 |
| Smoking initiation | 17 |
| Degree of unsaturation | 16 |
| Heel bone mineral density | 16 |
| Mean corpuscular volume | 15 |
| Neuroticism | 15 |
| Triglyceride levels | 15 |
| Mean corpuscular hemoglobin | 14 |
| Depression | 13 |
| COL2A1 protein levels | 11 |
| Prostate cancer | 11 |
| Short sleep duration (<5 hours) | 11 |
| Total fatty acid levels | 11 |
| Triglyceride levels (MTAG) | 11 |
| Depressed affect | 10 |
| Glycated hemoglobin levels | 10 |
| Hematocrit | 10 |
| High-density lipoprotein levels (MTAG) | 10 |
| Metabolic syndrome | 10 |
| Autism spectrum disorder or schizophrenia | 4 |
| **(B) 60 month** |  |
| **Trait** | **n** |
| CPVL protein levels | 13 |
| Type 2 diabetes | 12 |
| Attention deficit hyperactivity disorder or autism spectrum disorder or intelligence (pleiotropy) | 1 |
| Autism spectrum disorder | 1 |
| Cognitive aspects of educational attainment | 1 |
| General cognitive ability | 1 |
| Note: N represents the no of times the trait has been listed in the catalogue |  |

**Supplement table 2.** Sex- Interaction GWAS Trait frequency in (A) neonatal (B) 60 months.

|  |  |
| --- | --- |
| **(A) Neonatal** |  |
| **Trait** | **n** |
| Body mass index | 29 |
| Height | 12 |
| Neuroticism | 8 |
| Educational attainment | 6 |
| Externalizing behaviour (multivariate analysis) | 2 |
| **(B) 60 month** |  |
| **Trait** | **n** |
| Electrocardiogram morphology (amplitude at temporal datapoints) | 136 |
| Body mass index | 47 |
| Height | 40 |
| QT interval | 38 |
| SERPINA12 protein levels | 27 |
| SERPINA11 protein levels | 26 |
| Sex hormone-binding globulin levels | 26 |
| Pulse pressure | 22 |
| Blood protein levels | 20 |
| SERPINA9 protein levels | 18 |
| Systolic blood pressure | 18 |
| Hip circumference adjusted for BMI | 16 |
| Post bronchodilator FEV1/FVC ratio | 15 |
| SERPINA4 protein levels | 15 |
| Post bronchodilator FEV1 | 14 |
| CRTAC1 protein levels | 13 |
| Serum albumin levels | 12 |
| Albumin levels | 11 |
| Autism spectrum disorder or schizophrenia | 11 |
| Monocyte count | 11 |
| Insomnia | 10 |
| Low density lipoprotein cholesterol levels | 10 |
| Omega-6 fatty acid levels | 10 |
| Total testosterone levels | 10 |
| Type 2 diabetes | 10 |
| Cognitive function | 1 |
| Cognitive function (generalized correlation coefficient) | 1 |
| Cognitive function (immediate memory) (longitudinal) | 1 |
| Social communication problems | 1 |
| Note: N represents the no of times the trait has been listed in the catalogue |  |

**Supplement table 3.** Association of the significant CpGs with the subscale domains.

|  |  |  |
| --- | --- | --- |
| **Neonatal** |  |  |
| **Social Communication and Interaction (SCI)** |  |  |
| **CpG ID** | **$\beta$** | **p-value** |
| cg07132326 | -2.33431454 | 0.000114459 |
| cg09866104 | 2.496508785 | 0.000394318 |
| cg08848269 | -1.742057385 | 0.009330521 |
| cg25439867 | 1.982659869 | 0.002462201 |
| cg01439945 | -1.451178926 | 0.033756463 |
| cg10173649 | -1.578292563 | 0.0104598 |
| cg08670367 | 2.649579075 | 4.21541E-05 |
| cg22643307 | 3.143946903 | 2.81485E-06 |
| cg02193640 | 2.494241868 | 0.000188106 |
| cg04839673 | -1.520404894 | 0.021495225 |
| cg05708683 | -1.470622574 | 0.03549126 |
| cg12178267 | -1.998691146 | 0.003073319 |
| cg10389399 | -1.581202652 | 0.025016118 |
| cg06422883 | -1.674244317 | 0.013068806 |
| cg16187147 | -1.489176993 | 0.030412872 |
| cg13834633 | -1.903865305 | 0.002759814 |
| cg01180662 | 3.000760317 | 1.19084E-06 |
| cg04937388 | 2.250314139 | 0.00087109 |
| **Restricted and Repetitive Behaviors (RRB)** |  |  |
| cg07132326 | -1.373465467 | 0.01178406 |
| cg09866104 | 2.032157085 | 0.000896376 |
| cg25439867 | 2.133445038 | 0.000233633 |
| cg01439945 | -1.496949079 | 0.008426241 |
| cg08670367 | 2.160457496 | 0.000177475 |
| cg22643307 | 2.030900373 | 0.000699809 |
| cg02193640 | 1.841259665 | 0.001321538 |
| cg05708683 | -1.385945352 | 0.016805171 |
| cg16187147 | -1.213144382 | 0.036670208 |
| cg01180662 | 2.261057521 | 9.8257E-05 |
| cg04937388 | 1.215224816 | 0.034214828 |
| **5-year** |  |  |
| **Social Communication and Interaction (SCI)** |  |  |
| cg23860478 | -0.053966267 | 0.014337741 |
| cg04768606 | -0.05278272 | 0.002989862 |
| cg03353560 | -0.048074913 | 0.011667587 |
| cg11504923 | -0.076590846 | 0.000309715 |
| cg00486259 | 0.11453543 | 4.52E-09 |
| cg02040705 | -0.059584287 | 0.013825941 |
| **Restricted and Repetitive Behaviors (RRB)** |  |  |
| cg04768606 | -0.055879709 | 0.001579511 |
| cg11504923 | -0.057494164 | 0.003158848 |
| cg00486259 | 0.093315839 | 9.93E-07 |

